# A Logistic Model for Age-Specific COVID-19 Case-Fatality Rates

**DOI:** 10.1101/2020.06.12.20129908

**Authors:** Xiang Gao, Qunfeng Dong

**Author notes:** Corresponding author Qunfeng Dong, Center for Biomedical Informatics, Department of Medicine, Stritch School of Medicine, Loyola University Chicago, 2160 S. First Avenue, Maywood, Illinois 60153.

## Abstract

**Objectives:** To develop a mathematical model to characterize age-specific case-fatality rates (CFR) of COVID-19.

**Materials and Method:** Based on two large-scale Chinese and Italian CFR data, a logistic model was derived to provide quantitative insight on the dynamics between CFR and age.

**Results and Discussion:** We inferred that CFR increased faster in Italy than in China, as well as in females over males. In addition, while CFR increased with age, the rate of growth eventually slowed down, with a predicted theoretical upper limit for males (32%), females (21%), and the general population (23%).

**Conclusion:** Our logistic model provided quantitative insight on the dynamics of CFR.

**Lay Summary:** Recently published studies have qualitatively shown that the COVID-19 case-fatality rates increased with age, with elder people at higher risk of fatality than younger ones. In our study, we presented a quantitative mathematical modeling approach to re-analyze those published data. Specifically, we were able to derive a logistic model to characterize age-specific CFRs. The derived mathematical model uncovered novel quantitative insights on the dynamics between CFR and age. Specifically, we inferred from the model that while CFR increased with age, the rate of growth eventually slowed down, with a predicted theoretical upper limit of 23% for the general population.

## BACKGROUND AND SIGNIFICANCE

Recent studies showed that COVID-19 case-fatality rates (CFR) increased with age, with elder people at higher risk of fatality than younger ones^1,2,3^. Although those studies made important observations that COVID-19 CFR were age dependent, their results were limited by descriptive data analysis. In this study, we presented a quantitative mathematical modeling approach to re-analyze those published data. By deriving a logistic model to characterize age-specific CFR data, we were able to uncover novel quantitative insight on the dynamics of CFR.

## MATERIALS AND METHODS

### Data for mathematical modeling

The recently published Chinese and Italian CFR data were used for mathematical modeling since those were the only data with large sample sizes as of the date of the completion of our analysis on April 20, 2020: the Chinese data^1,3^ was compiled from 1023 case-fatality records as of February 11, 2020; the Italian data^2,3^ was assembled from 1625 case-fatality records as of March 17, 2020. These two data were used to compare our modeling results for the Chinese and Italian CFR data due to their similar sample size. In addition, a much larger Italian CFR data became available in a report^4^ dated March 26, 2020 to include gender-specific information for both females (4786 case-fatality records) and males (2010 case-fatality records). We used these gender-specific data to compare our modeling results for females and males. All the above CFR data were reported as the average CFR values for ages stratified by 10-year age groups (Table 1).

**Table 1.** Reported vs. Estimated CFR Stratified by Age.

| Age Range, year | China CFR, % |  | Age Range, y | Italy CFR, % |  | Italy Female CFR, % |  | Italy Male CFR, % |  |
| --- | --- | --- | --- | --- | --- | --- | --- | --- | --- |
|  | REP<sup>3</sup> | EST |  | REP<sup>2</sup> | EST | REP<sup>4</sup> | EST | REP<sup>4</sup> | EST |
| 0-9 | 0 | 0.01 | 0-9 | 0 | 0.00 | 0 | 0 | 0 | 0 |
| 10-19 | 0.2 | 0.02 | 10-19 | 0 | 0.00 | 0 | 0 | 0 | 0 |
| 20-29 | 0.2 | 0.05 | 20-29 | 0 | 0.00 | 0 | 0 | 0 | 0.01 |
| 30-39 | 0.2 | 0.15 | 30-39 | 0.3 | 0.03 | 0.1 | 0.015 | 0.6 | 0.051 |
| 40-49 | 0.4 | 0.47 | 40-49 | 0.4 | 0.14 | 0.4 | 0.10 | 1.1 | 0.29 |
| 50-59 | 1.3 | 1.29 | 50-59 | 1 | 0.78 | 0.8 | 0.63 | 2.4 | 1.61 |
| 60-69 | 3.6 | 3.61 | 60-69 | 3.5 | 3.83 | 3.5 | 3.55 | 6.9 | 7.46 |
| 70-79 | 8 | 7.99 | 70-79 | 12.5 | 12.23 | 11.6 | 11.99 | 19.8 | 20.34 |
| >=80 | 14.8 | 14.80 | 80-89 | 19.7 | 19.99 | 18.9 | 18.85 | 29.2 | 29.04 |
|  |  |  | >=90 | 22.7 | 22.54 | 20.4 | 20.65 | 30.8 | 31.37 |

REP: reported, EST: estimated

### Modeling procedure

For the modeling purpose, each reported CFR must be matched with a corresponding age. Since the reported CFRs were average values for people in each age group, we needed to estimate the average age for people in each age group. However, the age distributions in each group were not available in those published CFR reports. As an alternative, we estimated the age distributions for people in each age group based on the Chinese and Italian population-by-age data^5^ projected by the United Nation (UN) for ages stratified with 5-year age groups for the year 2020 (the closest year to date). To estimate the average age for people in a 10-year age group for the CFR data, a weighted arithmetic mean was computed with the two consecutive 5-year age groups in the UN population estimation. For example, the average age for people in the age group of 50-59 years equals the weighted arithmetic mean computed with the number of people of 50-54 and 55-59 years and their respective average ages in the UN population-by-age data.

After matching the reported average CFR with estimated average age in each age group, the nonlinear least-squares method was applied to fit logistic functions to model CFR with age. The logistic function has the form of *CFR* = *α/(*1*+ e*^*(γ - age)/β*^), in which *α, β*, and *γ* are model parameters and *e* is the natural logarithm base. The value of *age* and *CFR* are the input and output of the function, respectively. Using the R function *nls* with the option *SSlogis*, the expected values and standard errors of model parameters *α, β*, and *γ* were estimated from reported CFR data. The Kolomogorov-Smirnov test^6^ was applied compare the reported CFR values against the fitted logistic functions, using the function *ks*.*test* in R. Our computational code and data files are available at https://github.com/qunfengdong/COVID19-CFR.

## RESULTS AND DISCUSSION

### Model assessment

We found that a logistic model could characterize the mathematical relationship between CFR and age. Table 1 shows the comparison between the reported CFR in both Chinese and Italian data and the corresponding CFR values calculated using the fitted logistic model. Besides visual inspections (Fig. 1), the goodness-of-fit between the derived logistic functions and the reported CFR values also passed the Kolmogorov-Smirnov test (p=0.70 for the Chinese data, p=0.76 for the Italian data, including those for the females and males). It is worth mentioning that besides the Chinese and Italian CFR data, we were also able to fit the logistic model to a small CFR data^7^ of 44 case-fatality records from the United States (data not shown).

**Figure 1.**
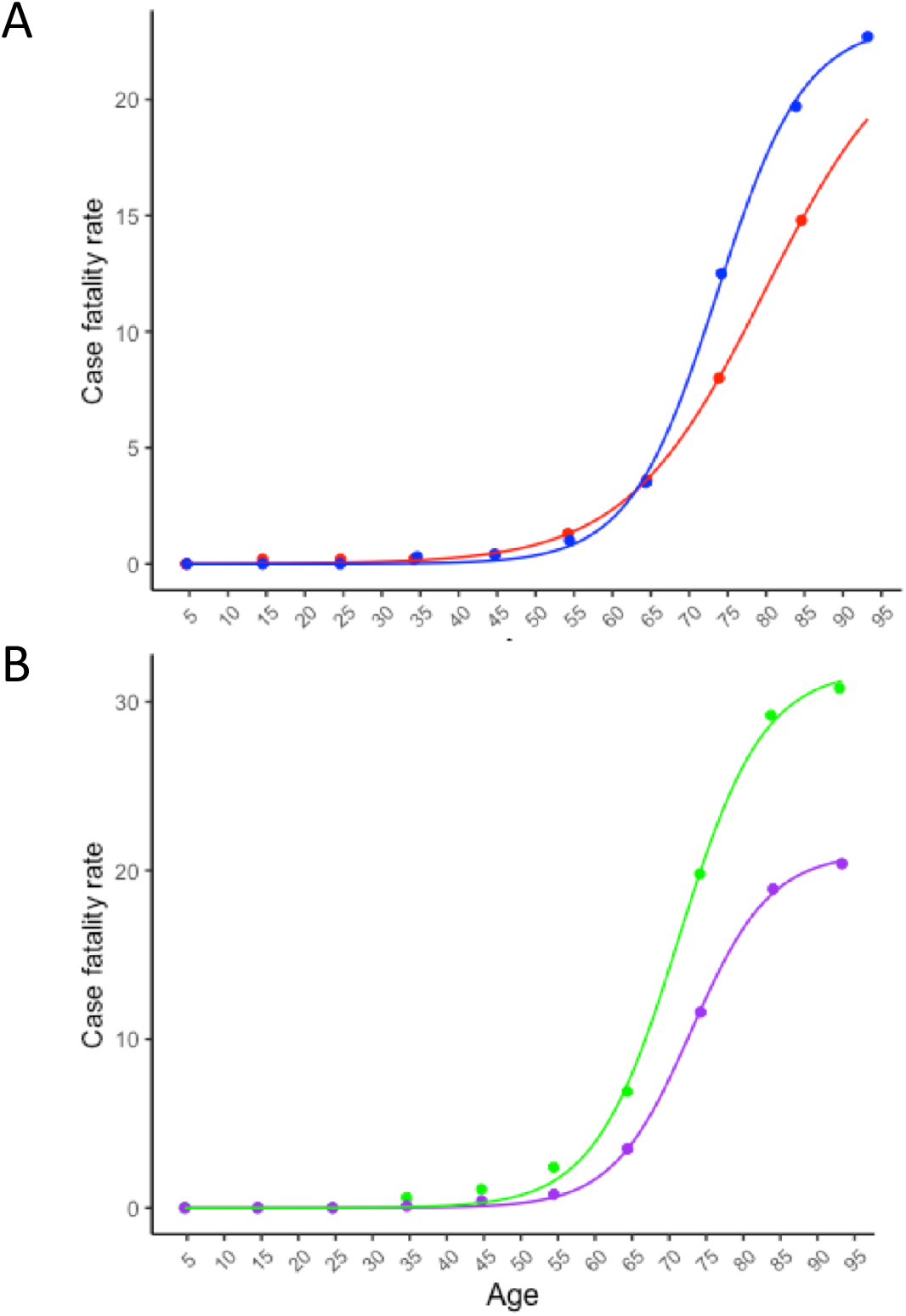
The fitted logistic function (lines) with the reported CFR data (dots) for **(A)** China (red) and Italy (blue), and **(B)** Female (purple) and male (green) in Italy. Table 1 displays the CFR data in tabular format.

### Dynamics of CFR

The fitted logistic model also characterized the dynamics between CFR and age, i.e., how exactly CFR increased with age, using three model parameters: *α, β*, and *γ*. Table 2 displays the values of those model parameters estimated from the reported CFR data.

**Table 2.** Estimated Logistic Parameters.

| **Data** | **$\alpha$** | **$\beta$** | **$\gamma$** |
| --- | --- | --- | --- |
| **China<sup>3</sup>** | 23.50±1.24 | 9.00±0.34 | 79.82±1.05 |
| **Italy (all gender)<sup>2</sup>** | 23.26±0.34 | 5.69±0.25 | 73.60±0.32 |
| **Italy (female)<sup>4</sup>** | 21.02±0.21 | 5.33±0.18 | 72.99±0.22 |
| **Italy (male)<sup>4</sup>** | 31.91±0.64 | 5.71±0.37 | 71.28±0.47 |

Mean ± standard error

The value of *α* corresponds to the theoretical upper growth limit for CFR in the logistic curve. Remarkably, *α* estimated from both the Chinese and Italian all-gender data were highly similar (i.e., 23.50% for China and 23.26% for Italy), indicating that while COVID-19 CFR increased with age, it had an upper limit around 23%. It is important to realize that 23% was the upper limit of CFR for the general population without specific gender consideration. The Chinese data did not include gender-specific CFR data, but the Italian data did. The estimated value of *α* was 21% and 32% for Italian females and males, respectively, indicating that males had a much higher CFR upper limit than females.

Since *β* represents the inverse of the maximum logistic growth rate, a smaller value of *β* corresponds to a faster growth rate during the exponential phase of the logistic curve. The estimated *β* for Italy (i.e., 5.7) was smaller than that of China (i.e., 9.0), indicating that CFR increased faster with age in Italy than in China. For example, CFR was 5.8% for the Chinese of age 70 and the Italian of age 67, and 17.5% for Chinese of age 90 and the Italian of age 80. In other words, for the same extent of increase of CFR from 5.8% to 17.5%, the age difference was 20 years for the Chinese and only 13 years for the Italian. Our modeling results also indicated that the CFR of Italian females (*β=*5.3) increased with age at the similar growth rate as Italian males (*β=*5.7).

Finally, *γ* corresponds to the age when the logistic growth rate of CFR is maximum. Our results indicated that while CFR increased with age, the rate of CFR growth started to decrease around the age of 80 years for Chinese, 74 years for Italian, 73 years for Italian females, and 71 years for Italian males.

### Calculate CFR for specific age

Although the COVID-19 CFR increased with age, the currently available age-dependent CFR information may not be suitable for personalized risk assessment due to the usage of arbitrarily stratified age groups. Both the Chinese and Italian CFR data were reported using age groups with 10-year ranges. Since the CFR data were reported as the average values for each age group, those reported values did not accurately reflect the risk of individuals of different ages within the same age group. For example, the Italian data showed that the average CFR for the age group of 60-69 years was 3.5% (Table 1). But a 69-year-old may have a very different CFR than a 60-year-old even though they belong to the same age group. Using the derived logistic model and the estimated parameter values in Table 2, CFRs can be calculated for specific ages instead of approximated with arbitrarily defined age groups. For example, using the aforementioned example of estimating the CFR for a 69.5-year-old Italian, we can plug in this specific age value (i.e., 69.5) into the logistic function derived from the Italian data, and obtain a CFR value of 5.67%. If this person were male, we can use the logistic function derived from the Italian male data, and obtain a CFR value of 7.62%.

## CONCLUSION

We developed a logistic model to characterize the reported COVID-19 age-dependent CFRs. The logistic model captures the observed trend that, as age increased, CFR initially increased slowly, and then increased rapidly before it slowed down again. The derived logistic model provided quantitative insight on the dynamics of CFR. Specifically, the model indicated that CFR increased faster in Italy than in China, yet with a similar growth rate between females and males. In addition, while CFR increased with age, the rate of growth eventually slowed down, and CFR gradually approach a theoretical upper limit. In addition, CFRs can be calculated for any specific age. In the future, the age-specific CFR calculated with our model may be further combined with other clinical data such as comorbidity for precise personalized risk assessment.

## Data Availability

We used published datasets that were referenced in our manuscript.

https://github.com/qunfengdong/COVID19-CFR

## Contributorship Statement

XG implemented the computational code, performed data analysis, and revised manuscript. QD conceived the project, performed data analysis, and drafted the manuscript.

